# COVID-19: Knowledge, Perception of Risk, Preparedness and Vaccine Acceptability among Healthcare Workers in Kenya

**DOI:** 10.1101/2021.10.19.21264712

**Authors:** Hafso Mohamed Abdulle, Moses Muia Masika, Julius Otieno Oyugi

**Author notes:** **Corresponding author** Hafso Mohamed Abdulle.

## Abstract

**Background:** Coronaviruses are highly contagious and healthcare workers are at a higher risk of contracting the disease. The objective of this study was to assess the level of knowledge, risk perception, preparedness for coronavirus disease 2019 and vaccine acceptability among healthcare workers in Kenya.

**Methods:** A cross-sectional study was conducted from December 2020 to January 2021. A link to an online self-administered questionnaire was disseminated to health workers across the country. SPSS version 20 was used for data analysis. Bivariate correlation analyses were used to determine associations between variables. P-value of <0.05 was considered statistically significant.

**Results:** A total of 997 participants were enrolled in the study. About half (53%) of the participants were female. The mean age was 36.54 years (SD = 8.31) and 46% of the participants were aged between 31-40years. The overall knowledge score of health workers for COVID-19 was 80%. Most of the health workers (89%) perceived that they were at high risk of infection. Seventy-two percent of the participants felt that they were either partially or fully prepared to handle patients with COVID-19. Overall, 71% of all health workers would take a vaccine if provided free by the government.

**Conclusion:** Health workers’ knowledge on transmission, clinical manifestations and risk factors for development of severe COVID-19 was good. Majority of the health workers perceived the risk of infection with COVID-19 as high and a significant number felt that they were not fully prepared to handle the pandemic. Majority of health workers would take a COVID-19 vaccine.

## Introduction

Coronaviruses cause infections that range from the common cold to severe acute respiratory syndrome (SARS). There are four genera in the coronaviridae family, which are alphaleto-, alpha- beta-, delta- and gamma-coronavirus (γ**-**CoV). α and β-coronaviruses have low pathogenicity hence similar to common cold since they present with mild respiratory symptoms [1]. MERS-CoV and SARS-CoV are two of the already known β-CoV which lead to respiratory infections that are severe and fatal.

In December 2019, in Wuhan, China, a number of cases of pneumonia of unknown origin were reported and a new strain of β-CoV was soon afterwards identified as the cause [2]. The genome sequence of this new β-coronavirus (SARS-CoV-2) that was isolated from a patient on 7 January 2020 was found to be 96.2% identical to SARS-CoV. It is thought that for humans to be infected, SARS-CoV-2 might have originated from bats to humans through intermediate hosts that are not known yet [1]. The disease caused by SARS-CoV-2 was on 11 February 2020 officially named as coronavirus disease 2019 (COVID-19) by the WHO [1]. To control the spread of COVID-19, WHO asked for a united effort of all countries as it declared on 30 January 2020 that the disease was a Public Health Emergency of International Concern (PHEIC) [3]. Transmission of COVID-19 from human to human occurs through droplets and direct contact and it may also occur via feco-oral route and this disease has an incubation period of 2 days to 2 weeks. For control of the COVID-19 infection, social distancing, wearing of masks, handwashing vaccine are some of the preventive measures that have been emphasized by WHO.

The COVID-19 pandemic is spreading at a very high rate. In 12 March 2021 minister of health declared the first COVID-19 case in Kenya. As of 2^nd^ August 2021, the confirmed number of COVID-19 cases was 198,234,951 while the deaths were 4,227,359 worldwide [5]. The African continent recorded a total of 4,917,071 COVID-19 confirmed cases and 117,304 deaths [6]. There was a total of 203,680 confirmed COVID-19 cases and 3,946 deaths reported by the government of Kenya [7] through the ministry of health.

COVID-19 has caused widespread panic and fear across the globe. Pure emotion of fear represents an individual’s removal from an immediate risk position [8]. Knowledge and attitudes of people influence the seriousness and extent of adherence of these people to the measures of personal protection and the clinical outcome. It is crucial that the COVID-19 clinical symptoms are known and well understood, this is even as the clinical symptoms are indicated as being nonspecific [9]. Fear of contracting a disease and even spreading it to respective families is reported by health workers most of the time [10]. This is based on the fact that healthcare workers (HCWs) are at a higher risk of exposure since some of the patients they handle may be infected, many without any symptoms [11]. A study done on health workers globally showed that the healthcare workers had insufficient knowledge on COVID-19 [12]. However, the study also showed that in terms of prevention of transmission of COVID-19, there were positive perceptions. To understand this disease, 33% of the HCWs involved in the study were found to rely on official government websites for information as their primary source. The level of knowledge of health workers was improved due to the updates given by the government [12]. In comparison, in terms of knowledge and practices, both the rural and urban health centers were found not to have considerable difference overall. However, health workers from urban areas were found to have scored slightly lower than their counterparts in precautionary practices such as doing a physical exam on suspected case [13].

Severity and likelihood are the two components that are involved in determining risk perception. Emotion such as concern and worry, and control illusions are some of the psychological components that can be used to influence and estimate perceived likelihood. A study in Hong Kong revealed that as new incidences and new cases were reported, more and more anxiety became evident in the community over time [14].

Kenya has taken measures to protect its citizens against COVID-19. Several interventions have been put in place to manage the infection rate including free testing for COVID-19, contact tracing, awareness campaigns, use of face-masks, social-distancing, regular hand-washing and vaccination among others. Despite these efforts leading to a reduction in COVID-19 spread, new COVID-19 cases are still reported every day. This study sought to assess the perceived risk of infection, level of knowledge of healthcare workers on clinical manifestation, transmission mechanism, treatment, COVID-19 vaccine and the rate of fatality as well as the perception of the level of preparedness for COVID-19 for individual healthcare workers and health institutions in the country. The study was conducted before the country began COVID-19 vaccination.

## Methods

### Study design and period

An online cross-sectional study was conducted to assess healthcare workers’ level of knowledge, perception of risk preparedness vaccine acceptability to handle COVID-19 in Kenya. It was conducted from December 2020 to January 2021.

### Study site and population

The study sites comprised of health facilities both the public and private hospitals throughout the country. We enrolled health workers (mainly medical doctors, nurses, clinical officers, and lab technologists) who work in private and public health facilities or medical training and research institutions in Kenya. Respondents were enrolled through convenience sampling technique.

### Variables

Dependent variables; these were used to measure or describe the problem that this study focuses on and they include:

➤ Level of Knowledge of measures on effective prevention
➤ Awareness of COVID-19 and knowledge about it
➤ Perception of risk

Independent variables – these were used to measure or describe factors which are assumed to affect the problem of study and they include:

➤ Demographics (age, sex, Profession, experience in years)
➤ County of work
➤ Type of facility (private, public, level of hospital, etc.)
➤ Any training on COVID-19

### Inclusion criteria

- Health workers (medical doctors, nurses, clinical officers, and lab technologists) based in Kenya
- Health workers who give consent for participation

### Sample Size

Using Fisher’s formula 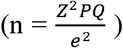 at a confidence level (1.96) for 95%, with; estimated prevalence (P) = 50%, our estimated the sample size was about 384 participants.

However, utilizing convenience sampling technique, we enrolled and collected data from a total sample size of 997 participants (health workers).

### Data Collection

Data was collected by use of a questionnaire that contained both open-ended and closed questions. We measured knowledge on COVID-19, perception of risk for infections with COVID-19 and preparedness of health workers to handle the pandemic. The questionnaire was administered online via REDCap. Health workers were sent a link to the questionnaire in a text message, social media and the email. There were 38 questions in the questionnaire where 35 of them were closed while the other 3 were open-ended. It had three sections – knowledge on COVID-19, risk perception, and preparedness to handle the COVID-19 pandemic. The knowledge section had questions on clinical features, COVID-19 vaccine and fatality rate. On the other hand, the risk perception section, explored the perceived risk to the general population, to health workers, self and family. The preparedness section had questions on training on COVID-19 and infection control, supplies, stigmatization of health workers who had recovered from COVID-19 and confidence of health workers in their ability to manage a COVID-19 patient. The data collected from the participants on REDCap application was transferred to the IBM SPSS Statistics software version 20 for analysis.

### Data management

IBM SPSS Statistics software was utilized for data analysis. Descriptive characteristics are presented in text, table and graphs. We used chi-square to test for association between categorical variables and t-test to measure association between numerical and categorical variables. Medical doctors, pharmacists and dentists were analyzed in the same category as doctors.

Nairobi city and Mombasa counties were categorized as urban and the rest of the counties were categorized as rural or semi-rural. A p value of ≤ 0.05 was considered statistically significant.

### Ethical considerations

We sought for approval to conduct the study from UoN-KNH Ethics and Research Committee before sharing the consent form and the questionnaire to collect data from the respondents. Once the study was approved (**P338/06/2020**), participants were provided with informed consent form through an online link on the REDCap application.

## Results

### Study participants Characteristics

Nine hundred and ninety-seven health workers were enrolled from all 47 counties in Kenya. The proportion and gender of the respondents per cadre is shown in **Fig 1**. The mean age of all respondents was 36.54 years (SD = 8.31). The age ranged from 22 to 78 years with the largest age group being 31 – 40 years (50.9%). For more respondents’ characteristics see **Table 1**.

**Figure 1:**
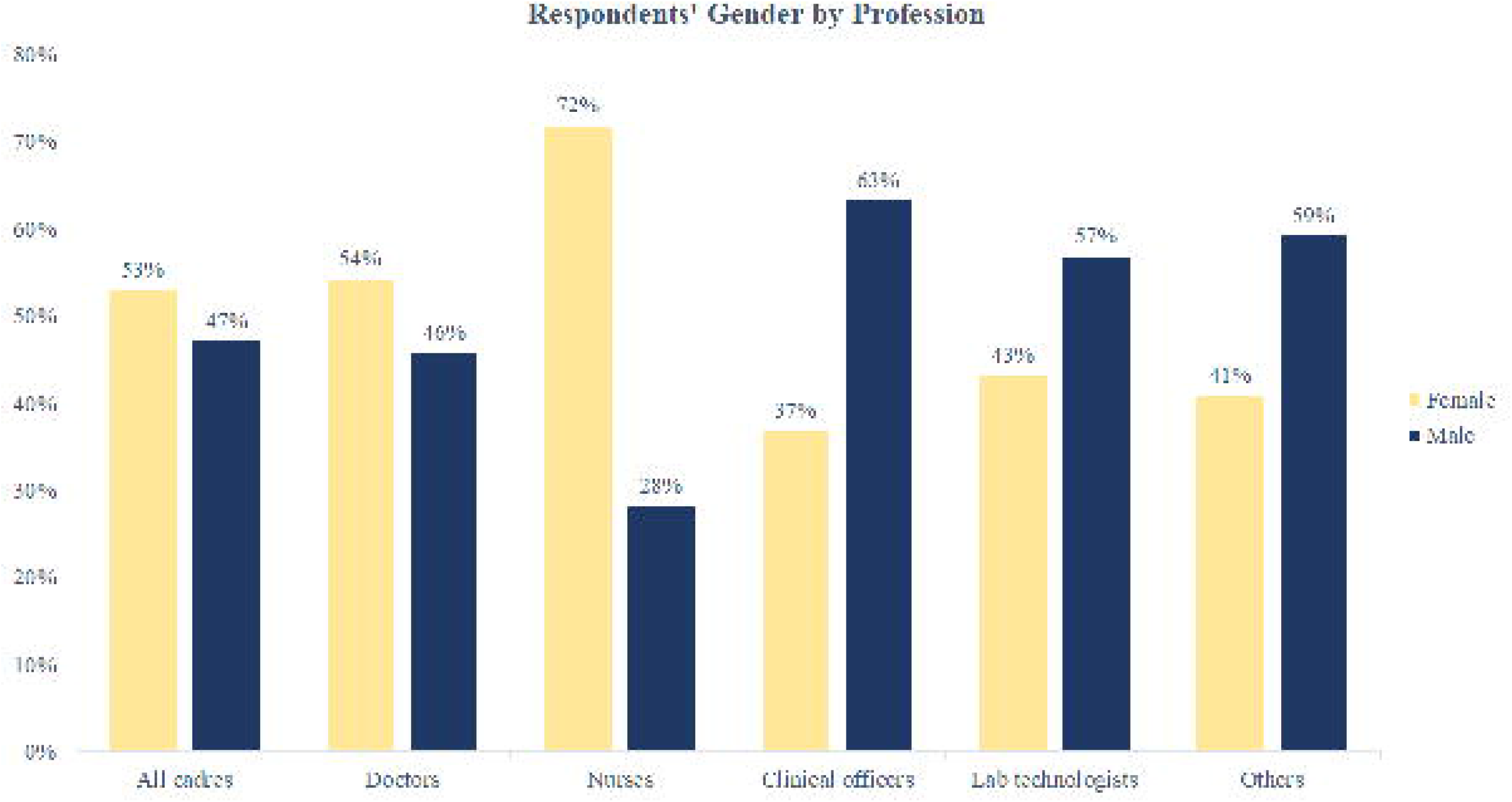
Gender proportions by health profession

**Table 1:** Respondents’ characteristics.

| Table 1: Respondents' Characteristics |  |  |
| --- | --- | --- |
| Parameter | Groups | n (%) |
| Gender | Male | 469 (47%) |
|  | Female | 526 (53%) |
| Age |  |  |
| Mean | 36.54 |  |
| SD | 8.31 |  |
| Median | 35.00 |  |
| IQR | 10 |  |
| Range(Min-Max) | 22-78 |  |
| Age groups | 22-30 years | 226 (23%) |
|  | 31-40 years | 458 (46%) |
|  | 41-50 years | 150 (15%) |
|  | 51 and above years | 66 (7%) |
| Profession | Doctors | 344 (34%) |
|  | Nurses | 262 (26%) |
|  | Clinical officers | 209 (21%) |
|  | Lab technologists | 118 (12%) |
|  | Others | 73 (7%) |
| Place of work | National hospital | 143 (14%) |
|  | County facility | 541 (54%) |
|  | Private facility | 160 (16%) |
|  | Training and research institution | 81 (8%) |
|  | Faith-based facility | 36 (4%) |
|  | Others | 40 (4%) |
| Level of education | Certificate | 17 (2%) |
|  | Diploma | 336 (34%) |
|  | Bachelor's degree | 390 (39%) |
|  | Post-graduate qualification | 248 (25%) |
| Work experience | Less than 2 years | 72 (7%) |
|  | 2-5 years | 184 (19%) |
|  | 6-10 years | 261 (26%) |
|  | 11-20 years | 277 (28%) |
|  | More than 20 years | 193 (19%) |
| County | Urban | 323 (32%) |
|  | Rural and semi-rural | 645 (65%) |
| Working in a COVID-19 isolation facility | Yes | 206 (20%) |
|  | No | 778 (78%) |
| Frequency of working in a COVID-19 isolation or testing facility | Every day | 107 (11%) |
|  | 2 to 3 times a week | 50 (5%) |
|  | Once a week or less | 47 (5%) |
| Live alone | Yes | 140 (14%) |
|  | No | 848 (85%) |

### Level of knowledge of HCWs for COVID-19

Knowledge score was compared across different professional cadres. There was a statistically significant difference in knowledge score across different groups. The results from these tests are shown in **Table 2**.

**Table 2:**
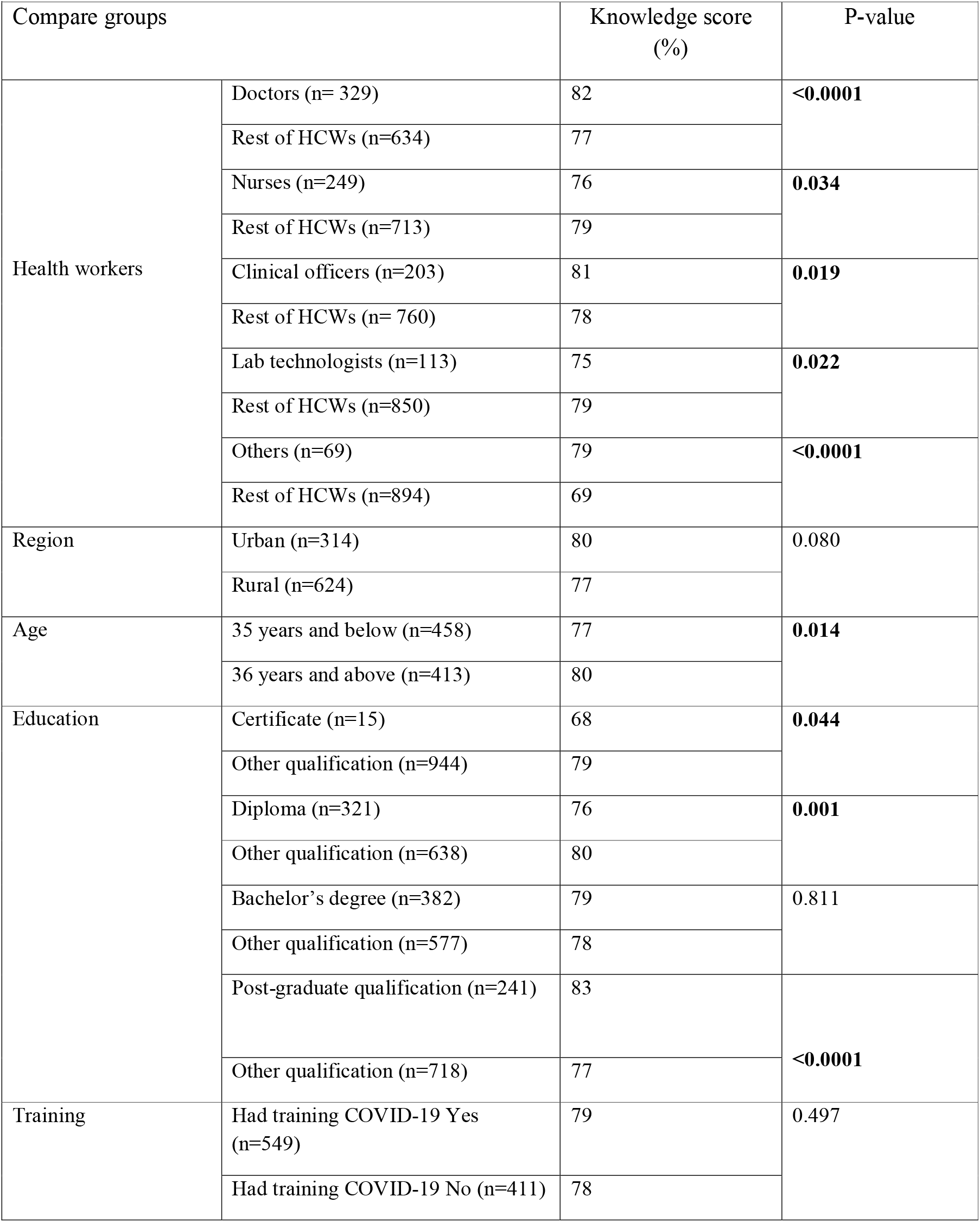
Knowledge score on COVID-19 among health workers.

Most (94%) of all the respondents identified fever as a symptom of COVID-19. Refer to **Fig 2** for the score for each symptom. The majority of HCWs were able to identify key risk factors for severe COVID-19 such as diabetes (93.8%), heart disease (84.4%), and cancer (77.5%). Meanwhile, (12.6%) of the respondents wrongly identified young age as a risk factor for development of severe COVID-19.

**Figure 2:**
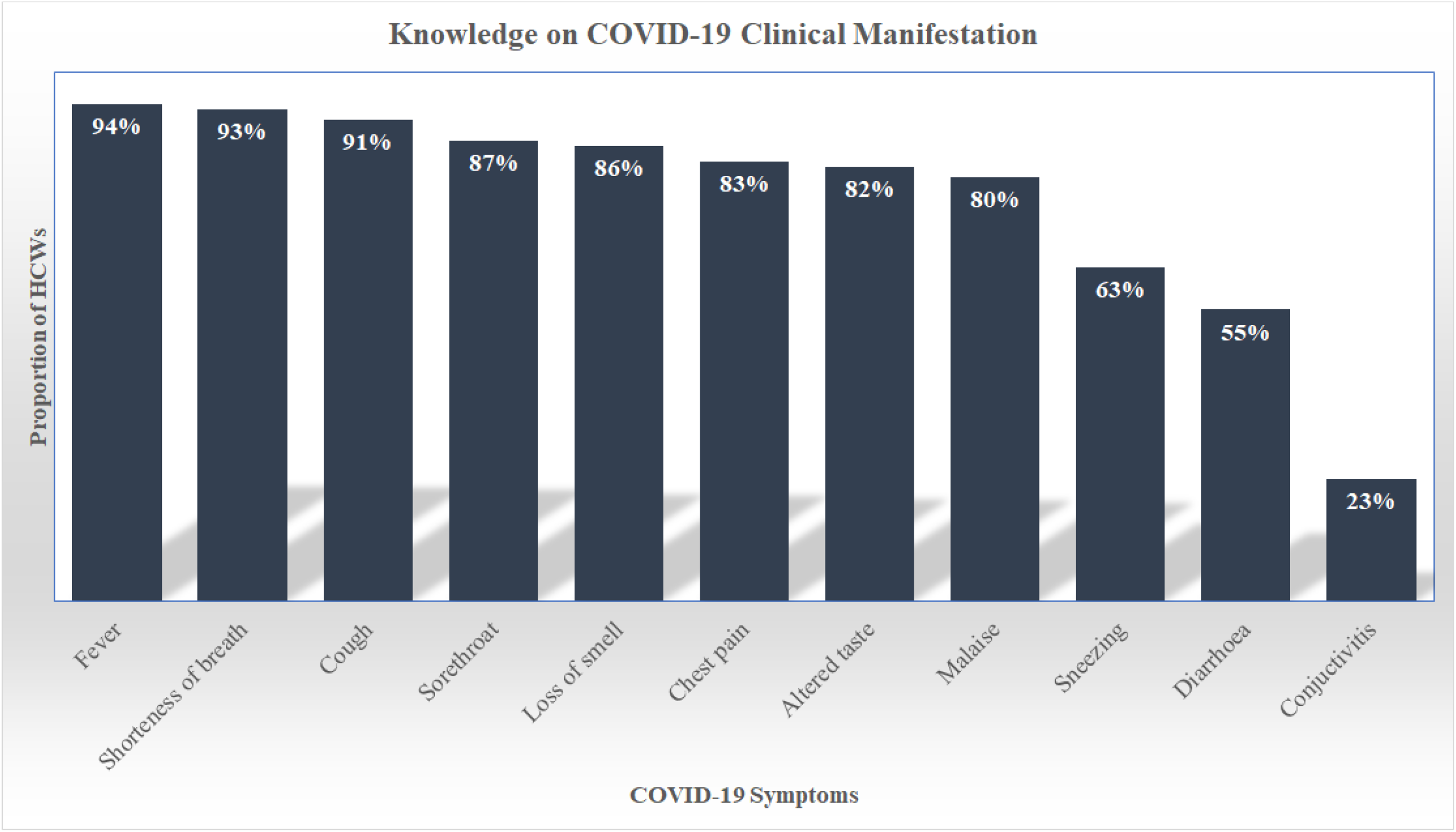
Health workers’ knowledge on clinical manifestations for COVID-19

### Perception of risk of infection with COVID-19 for HCWs

A majority (88%) of the respondents (n=997) perceived the level of risk of infection for the general public as high while 9% of the HCWs perceived it as low. Compared to the rest of the cadres, nurses were more likely to report the level of risk of infection for COVID-19 to general public as high (P=0.001). Doctors had the lowest number of respondents (85%) who felt that the level of risk for the general public getting infected with COVID-19 was high (P<0.001). Ninety-seven percent of nurses perceived the level of risk of contracting COVID-19 among health workers to be high as did 94% of the lab technologists. There was no significant relationship between the perceived risk of getting a COVID-19 infection for HCWs and different health cadres.

Most nurses and lab technologists felt that there was a high level of risk of infection with COVID-19 for individuals (self). There was a significant relationship between perception of risk of COVID-19 infection for self and nurses (94%, P=0.006) and with lab technologists (80%, P=0.005).

A majority (91%) of the healthcare workers indicated that they would be comfortable to work with a colleague who had recovered fromCOVID-19. This ranged from 86% for ‘other cadres’ to 96% for doctors

### Preparedness for COVID-19 among Health Workers

A total of 673 (70%) of health workers reported that they had received training on COVID-19. Doctors were more likely to have been trained on COVID-19 than other cadres (75% vs 64%, P<0.001). See **Table 3**.

**Table 3:**
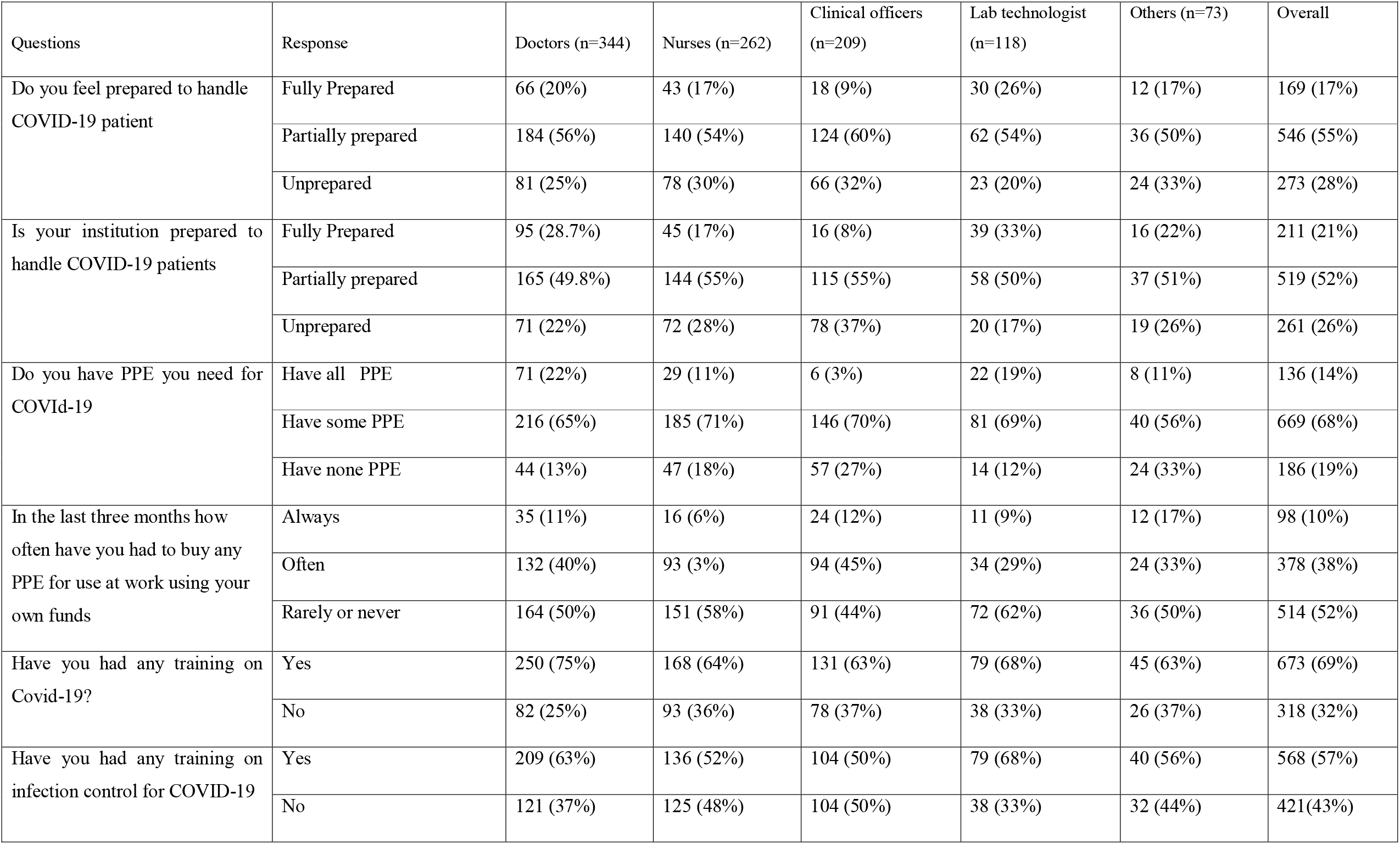
Perception of the Level of Preparedness for COVID-19 against healthcare Cadre.

A third (32.1%) of health workers from private facilities felt that they were fully prepared to handle patients with COVID-19 as compared to 11.9% of health workers from national hospitals who felt that they were fully prepared. On the other hand, private facilities (40.9%) were reported to be prepared to handle patients with COVID-19 while only 11.5% of the national hospitals were reported to be fully prepared.

Sixty-eight percent of the respondents indicated that they received training on COVID-19. While private facilities reported the highest number (78%) of HCWs who received a COVID-19 training, 62.7% of respondents from national hospitals reported that they had received training on COVID-19 infection. A chi-square test revealed that there was a significant relationship between place of work (health facility) and whether one got training for COVID-19 or not (P=0.019).

Most respondents from urban counties (64%) reported that they had received training specifically for COVID-19 infection control as opposed to respondents from rural and semi-rural counties (54%) who reported that they received this kind of training. There was a significant association between the location of the counties in which the HCWs work and having had training specifically for COVID-19 infection control (P=0.003).

Respondents (35%) from urban counties reported that their facilities were fully prepared to handle COVID-19 patients while 15% of respondents from rural and semi-rural counties felt they were fully prepared. There was a significant relationship between county location and its facility’s preparedness to handle patients with COVID-19. Health facilities in urban counties were more likely to be prepared to handle COVID-19 patients as compared to facilities in rural and semi-rural counties (P<0.001).

### Acceptability of COVID-19 vaccine among HCWs professions

Overall, 71% of all healthcare workers would take a COVID-19 vaccine if provided free by the government. Doctors were most likely (76%) while nurses were least likely (64%) to accept the vaccine. There was a significant relationship between professional cadre and acceptability of COVID-19 vaccine as shown in **Table 4**. Vaccine acceptability for each cadre is shown in **Fig 3**.

**Table 4:** Vaccine Acceptability among Healthcare Workers.

| **Parameters** | **Groups** | **Vaccine acceptability** | **P-value** |
| --- | --- | --- | --- |
| Gender | Male (n=460) | 77% | <0.0001 |
|  | Female (n=513) | 66% |  |
| Age | Below 35 years (n=347) | 74.8% | 0.004 |
|  | Above 36years (n=276) | 66% |  |
| County of work | Nairobi (n=272) | 70% | 0.78 |
|  | Other counties (n=674) | 71% |  |
| Doctor | (n=330) | 76% | 0.01 |
| Others | (n=643) | 68% |  |
| Nurses | (n=252) | 64% | 0.007 |
| Others | (n=721) | 73% |  |
| Previous infected COVID-19 | Yes (n=56) | 73% | 0.7 |
|  | No (n=918) | 71% |  |
| Work at a COVID-19 treatment centre or testing lab | Yes (n=201) | 62% | 0.002 |
|  | No (n=763) | 73% |  |
| Live alone | Yes (n=135) | 77% | 0.08 |
|  | No (832) | 70% |  |

**Figure 3:**
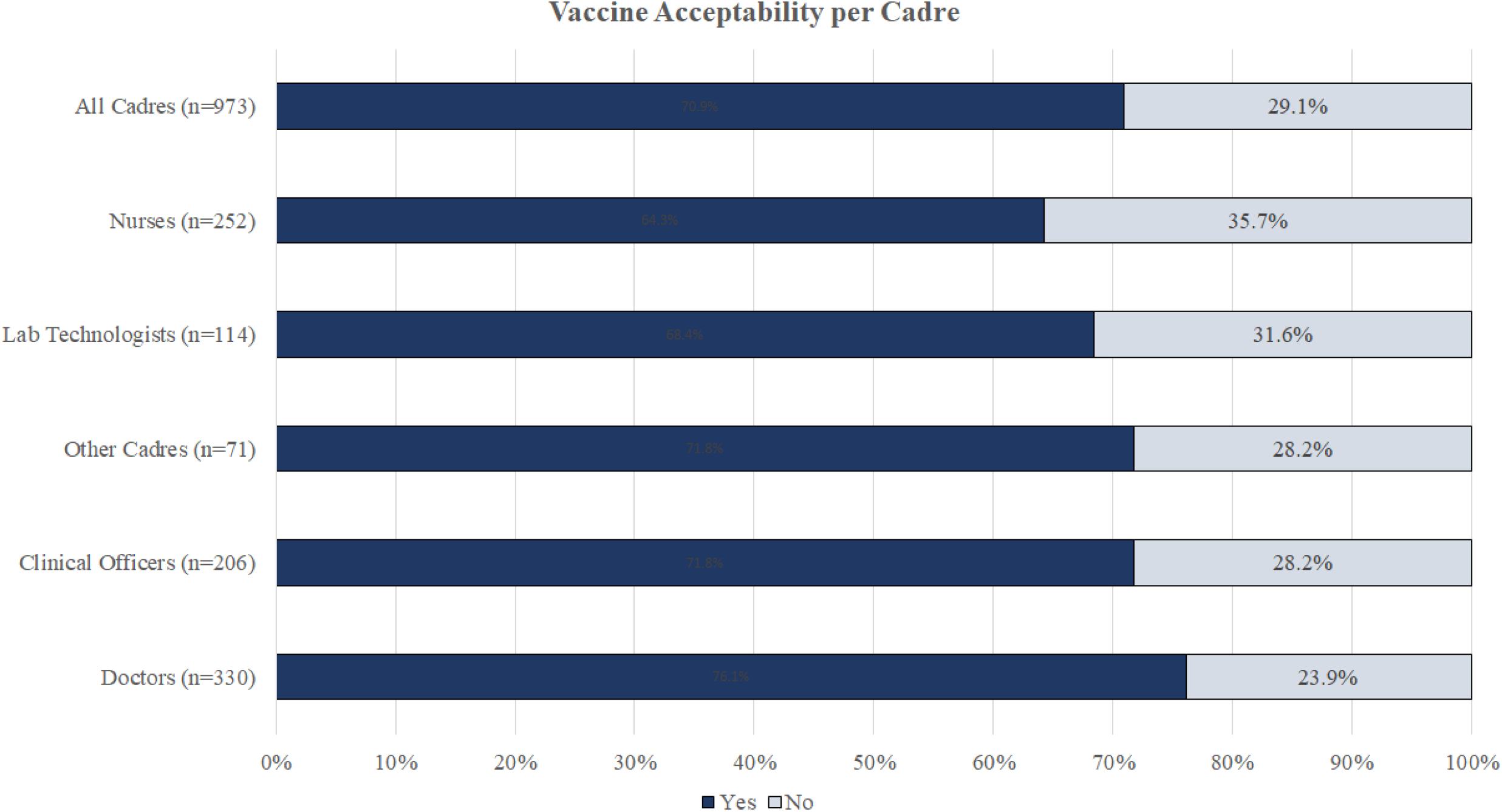
Proportion of health workers who would take a COVID-19 vaccine

### Concerns of Health Workers regarding COVID-19

Participants highlighted their main concerns about COVID-19. A majority of the participants were more concerned about getting infected with the disease and then in-turn passing it to their family members or colleagues. This is especially if their family members or colleagues had a chronic disease that may lead to a medical complication. Some of the health workers were worried about the inadequacy of personal protective equipment in order to enhance their safety while others were concerned about the ignorance displayed on measures put in place to control the infection and how generally naïve the public is in regards to risks associated with COVID-19.

These are some of the comments by the respondents:

“Getting infected…Infecting family members” a nurse (41-50 years).

“I am asthmatic and hypertensive. My age also puts me at risk” a clinical officer (51-60 years).

“Ignorance among the population on containment measures they don’t fully observe (prevention measures)” a clinical officer (31-40 years).

### Facilities’ preparedness for COVID-19

In order to control and curb the spread of the virus, participants felt there was a need to train more HCWs on COVID-19 infection control, create more awareness and sensitize both the health staff and the general public among other measures. A majority of the respondents in this group felt that in order for their health institutions to be prepared to handle COVID-19, a number of things had to be improved – hiring of more health workers, procurement of additional medical equipment such as ventilators, availing more PPEs, installation of oxygen plants at the hospitals, setting up well-equipped isolation facilities where there were none and expanding the existing COVID-19 isolation facilities where they existed.

“To stock full PPE kit, to put up a functional ICU, motivate health workers and enhance COVID-19 training” a clinical officer (31-40 years).

### Reasons for declining a COVID-19 vaccine

Reasons for declining a COVID-19 vaccine were mainly concerns about the efficacy, unknown side effects and safety of the vaccine. While other respondents felt that vaccines were developed as a monetary scheme, some felt that the vaccine developed might not be safe for use due to the “hurried” approval or that it may not be effective due to the mutative nature of the virus. While other respondents cited lack of trust for their government and politicians as reason for vaccine hesitancy, other healthcare workers felt that they did not have enough information or knowledge on the COVID-19 vaccine and require more information on the vaccine before using it. Others indicated that they would rely on herd immunity and therefore did not need to take the vaccine. Respondents’ comments on these concerns were as follows:

“I fear the side effects of the (COVID-19) vaccine” a nurse (31-40 years).

“I don’t trust this government’s effort in this fight against COVID-19. I may just get infected after the vaccination due to hidden agendas” a clinical officer (31-40 years).

## Discussion

It is critical that health facilities and most importantly health workers are prepared to handle any highly infectious disease at any given time. This will enable them to protect themselves and the general public against such infections. The objective of this study was to determine the level of knowledge, perception of risk and the preparedness of HCWs for COVID-19 in Kenya.

In our study, forty-seven percent of the participants were male, the mean age was 36.54 years (SD=8.31) and 64% of the participants had at least a bachelors’ degree. In another study on Turkish health workers, 27% of the participants were male while the participants mean age was 33.88 years (SD=8.72) and 97.6% indicated they were university graduates. Though the level of knowledge on COVID-19 was good, it differed between the two studies (80% vs 91.7%). The difference in level of education attained by health workers in the two studies may be responsible for the different levels of knowledge on COVID-19 between the two groups (countries).

A majority of the respondents had knowledge on the risk factors and clinical manifestation of COVID-19 with at least 63% identifying the symptoms of the disease. Our findings were in tandem with some of the other similar studies done in other countries. A similar study in Northwest Ethiopia reported that 73.8% of the health workers had good knowledge on COVID-19 [15]. Our finding was lower than that in Ethiopia. In contrast, a study on dental practitioners from 23 countries across the world reported a lower mean knowledge score of 34.9% [16]. The possible reason for the difference in the knowledge scores between these studies may be due to the different settings for healthcare in these countries. Time may have also played a role in the difference due to more awareness on COVID-19 being created. Knowledge on key risk factors associated with development of severe COVID-19 was also found to be high. However, a proportion of the respondents (12.6%) wrongly highlighted that ‘young age’ was one of the risk factors that was associated with severe COVID-19 development.

Overall, level of perception of risk of infection with COVID-19 for health workers was found to be high (97%). Respondents also felt that the risk of infection with COVID-19 for the general public was also high (90%). In another study on the Turkish health workers, 69.32% of the respondents perceived the infection with COVID-19 as dangerous [17]. The sense of preparedness could be the reason for the difference between the two studies and therefore might explain why the nurses perceived the risk of infection with COVID-19 as high for everyone.

Most health workers would be comfortable to work with a colleague who had recovered from COVID-19. This could be associated with the good knowledge score of the participants on the modes of transmission, and the level of education attained by an individual like a health worker having attained at least a bachelors’ degree.

Of all the participants who took part in the study, a majority felt at least partially prepared (72%) to handle the COVID-19 pandemic. However, some gaps such as inadequate PPE and training for COVID-19 infection control were identified. In comparison with a study in Ethiopia, the participants who reported that they were prepared for COVID-19 were lower (59.5%) [18] than in our study. Health workers may feel a sense of preparedness to handle a pandemic when they are trained adequately and provided with the necessary PPE. This may in-turn lead to change of attitude towards improved health service delivery. Inadequate PPE and lack of training are likely to put the health workers at a risk of contracting the infection. This in turn affects the expected service delivery and therefore complicates the fight against COVID-19. While more doctors and lab technologists felt that their institutions were fully prepared to handle patients with COVID-19, only 8% of the clinical officers felt the same leaving room for exploring why this difference in particular exists.

There was a difference between the urban counties and the rural and semi-rural counties in terms of preparedness for control the COVID-19 pandemic. Sixty-four percent of the participants from the urban counties indicated that they had received training specifically for COVID-19 infection control unlike their colleagues from the rural and semi-rural counties that were accounted for by 54.1%. This shows that there are factors that could contribute to such an occurrence. Since there are often fewer and isolated health workers in the rural and semi-rural counties as compared to their counterparts in the urban counties, the reasons for the difference in level of knowledge and preparedness to handle the pandemic might be due to poor connectivity and poor networking among health workers as well as government policies that are urban centered.

A significant number of the respondents (29%) would not take a COVID-19 vaccine if one was offered at the time of the study. Most of the reasons such as possible side effects, efficacy and the ‘hurried vaccine approval’ cited for the hesitancy can be addressed by providing the correct information on Covid-19 vaccines. In a similar study done in the United States, the reasons for vaccine hesitancy among healthcare workers included safety concerns, the approval speed and effectiveness, however, only 36% of the participants would take the vaccine [19]. The concern on the efficacy of vaccines (Oxford–AstraZeneca) under clinical trial have been explained, in that, these vaccines reportedly have more than 90% efficacy against COVID-19 [20].

From our study it was determined that male health workers were more likely to accept the COVID-19 vaccine than the female (77% vs 66%). In comparison to a similar study in Saudi Arabia, it was also found that male health workers were more likely to take a COVID-19 vaccine unlike their female counterparts (67% vs 33%) [21]. From the two studies, being a female health worker lowered the probability of taking a COVID-19 vaccine.

A few limitations were found in this study; some healthcare workers who participated in the research study did not fully fill the questionnaire. The questionnaire was made as short as possible and by using a user-friendly design hence a response rate of 95%. There also may have been a selection bias by leaving out healthcare workers who may not have had access to internet. Another limitation is that the REDCap application is configured for ease of use on a smart phone or a computer and therefore health workers without one of these devices may have been unable to participate. This is an inherent weakness in the study.

## Conclusion

Knowledge of healthcare workers on clinical manifestations and risk factors for development of severe COVID-19 was good. With good knowledge, the health workers are in a good position to manage and control the spread of COVID-19. However, a majority of the HCWs perceived the risk of infection with COVID-19 as high and a significant number of them felt that they were not fully prepared to handle the pandemic. The perception of high risk of infection as well as feeling of being unprepared can affect the psychological well-being of the health workers, thus, affecting their service delivery. A majority of health workers (about two thirds) were willing to take a COVID-19 vaccine if provided with one. COVID-19 vaccination was introduced in Kenya after this study was conducted; we therefore recommend another study on vaccine uptake and its facilitators or barriers among healthcare workers in the country.

## Recommendation

There is need to fill the knowledge gap among different health cadres to avoid the differences observed. There is also need for robust training on COVID-19 infection control and provision of PPE to boost knowledge and enhance preparedness and improve perception of risk of infection with the disease.

## What is already known about this topic

- COVID-19 is a highly contagious disease and healthcare workers are at a higher risk of contracting the disease.
- Knowledge, attitudes and preparedness of people influence the seriousness and extent of adherence of these people to the measures of personal protection and the clinical outcome on emergent diseases or infections.
- Researchers have developed a number of vaccines to help prevent COVID-19 from spreading further.

## What this study adds

- The findings from this study provide an insight on how knowledge, perception of risk and preparedness for COVID-19 have impacted on health workers and how this may affect health service delivery.
- The findings from this study are important as they can be used in the future pandemics in addressing the hesitancy of vaccine uptake.

## Data Availability

All data produced in this study are contained in the manuscript and its supplemental files.

## Authors’ contributions

Hafso Mohamed and Moses Masika: conceptualization of the study, wrote the introduction, results; Hafso Mohamed, Julius Oyugi and Moses Masika: data collection, discussion of results and wrote the conclusion. All the authors read and agreed to the final manuscript.

## Acknowledgements

We deeply appreciate Collins Mwebi Maina and Saadia Ahmed Guhad for their assistance and commitment in drafting this manuscript. Moses Masika would like to acknowledge that he is supported by a career development fellowship grant from the European and Developing Countries Clinical Trials Partnership (EDCTP TMA2017CDF-1865)

