## Supplemental Questionnaire for "COVID-19: Knowledge, Perception of Risk, Preparedness and Vaccine Acceptability among Healthcare Workers in Kenya" (from another version of this preprint: version 1, 2021-10-22)

**1A)**

### **Study Questionnaire**

Investigators: **Dr. Hafso Mohamed Abdulle, Dr. Moses Masika, Prof. Julius Oyugi**

University of Nairobi Institute of Tropical and Infectious Diseases (UNITID)

1. What is your Profession?

- Medical Doctor
- Dentist
- Pharmacist
- Nurse
- Clinical Officer
- Lab technologist
- Public Health Officer
- Other:

2. What is your place of work?

- National Hospital
- County Hospital
- Sub-county Hospital
- Health centre/dispensary
- Private Hospital
- Private Clinic
- Training institution
- Other:

3. Which County do you work in?	*(drop down list)*

4. Do you work in a Covid isolation facility or Covid testing Lab?	Yes	No

5. How many times do you work in a Covid Isolation facility or testing Lab?

- Everyday
- 2 to 3 times a week
- Once a week
- Less often than once a week

6. What is your gender:	Male	Female

7. What is your age:

8. How long is your work experience?

- Less than 2 years
- 2 – 5 years
- 6 – 10 years
- 11 – 20 years
- More than 20 years

9. What is the risk of infection with Covid-19 for the average Kenyan?

- Very high
- High
- Low
- Very low
- I don’t know

10. What is the risk of infection with Covid-19 for health care workers in Kenya?

- Very high
- High
- Low
- Very low
- I don’t know

11. What is the risk of infection with Covid-19 for you?

- Very high
- High
- Low
- Very low
- I don’t know

12. Do you live alone?	YES	NO

13. Are you concerned that you may infect your family members with Covid-19?

- Always concerned
- Very often concerned
- Sometimes concerned
- Rarely concerned
- Never concerned

14. What is/are your main concerns about Covid-19?

*Text box*

15. What is the fatality rate of Covid-19?

- Less than 1%
- 2 – 10%
- 20 – 50%
- Over 50%

16. What are the clinical manifestations of COVID-19? (Tick all that apply)

- Fever
- Cough
- Malaise
- Sorethroat
- Sneezing
- Shortness of breath
- Diarrhoea
- Chest pain
- Ear ache
- Skin rash
- Insomnia
- Loss of smell
- Altered taste
- Conjunctivitis

17. When is the infection with Covid-19 mostly likely to occur during?

- While putting on PPE
- While attending to Covid-19 patients in full PPE
- While taking off PPE after work

18. Do you feel prepared to handle a patient with Covid-19?

- Prepared
- Partially prepared
- Unprepared

19. Is your facility/institution prepared to handle covid-19 patients?

- Prepared
- Partially prepared
- Unprepared

20. Do you have the personal protective equipment (PPE) you need for Covid-19?

- I have all the PPE I need
- I have some PPE
- I have no PPE

21. In the last three months, how often have you had to buy any PPE for use at work using your own funds?

- Always
- Very often
- Often
- Rarely
- Never

22. Have you had any training on Covid-19?	YES	NO

23. Have you had any training specifically for infection control for COVID-19?	YES	NO

24. Would be comfortable to work with a colleague who has recovered from Covid-19?	YES	NO

25. What more do you think your facility/institution needs to do to prepare for Covid-19?

*Text box*

26. Would you accept a COVID-19 Vaccine if the government provides one for all Healthcare workers? Yes No

27. Please indicate your reason for declining the vaccine

*Text box*

28. Which conditions are associated with a high risk for severe Covid-19 (Check all that apply)

- Diabetes
- Heart disease
- Young age
- HIV5
- Cancer

29. Have you been infected with Covid-19? (Laboratory confirmed infection)	Yes	No
